# Structured Occlusion Reveals State-Dependent Smooth Pursuit Deficits Across Acute and Chronic Psychosis

**DOI:** 10.64898/2026.07.03.26357245

**Authors:** Tzofia Simkovich, Ilan Segal, Yoram S. Bonneh, David Israeli

**Author notes:** Yoram S. Bonneh and David Israeli jointly supervised this work and contributed equally as senior authors. **Correspondence:** Yoram S. Bonneh. **Clinical correspondence:** David Israeli.

## Abstract

Smooth pursuit eye movement abnormalities are well established in psychosis, but the specific components of pursuit performance that vary across clinical states and symptom profiles remain insufficiently characterized. Here, we used a rapid smooth pursuit paradigm combining standard linear tracking (repeated short trials moving in different directions) with structured target occlusion to examine oculomotor performance in individuals with acute psychosis, chronic psychosis, and healthy controls. The occlusion condition allowed assessment of tracking when the target was temporarily hidden and gaze had to be maintained along the expected trajectory. Basic oculomotor measures, including full pursuit gain and initial catch-up saccade properties, were largely preserved in patients. In contrast, more specific trajectory-based measures revealed distinct abnormalities. Saccade-free smooth tracking gain was selectively reduced in acute psychosis, whereas tracking deviation during non-occluded pursuit was altered in both patient groups, reflecting reduced forward tracking relative to controls. During structured occlusion, patients showed reduced forward gaze progression along the expected trajectory, with a graded pattern across groups: controls showed the strongest predictive lead, chronic patients an intermediate response, and acute patients the weakest predictive lead. Tracking deviation and occlusion-related deviation were both associated with positive symptom severity, whereas smooth tracking gain was not. These findings suggest that smooth pursuit abnormalities in psychosis are measure-specific rather than uniform, involving both broader psychosis-related alterations in gaze–target alignment and state-sensitive disruptions in occlusion-related tracking. Structured occlusion may therefore offer a useful extension of conventional smooth pursuit paradigms for probing prediction-related sensorimotor control in psychosis and distinguishing acute from chronic clinical states.

## Introduction

Far from being mechanical adjustments of gaze, eye movements reflect the brain’s ability to coordinate sensory input, motor planning, and cognitive processes in real time (Spering & Montagnini, 2011; Kadosh et al., 2024; Rizzuto et al., 2026; Rosenzweig & Bonneh, 2020; Laubrock & Kliegl, 2015; Hayes & Henderson, 2017), making them a useful assay for studying altered sensorimotor and cognitive function. Among oculomotor behaviors, smooth pursuit eye movement (SPEM) is particularly relevant because it requires continuous tracking of a moving target and ongoing coordination between visual input and motor control.

Smooth pursuit eye movement is a continuous oculomotor behavior that allows the eyes to follow a moving target while keeping its image aligned with the fovea and preserving visual clarity throughout motion. Smooth pursuit requires the integration of visual motion input with motor commands and must compensate for inherent visuomotor delays (Lisberger et al., 1987; Barnes, 2008). This compensation is not achieved solely through retinal error signals, which reflect discrepancies between target and eye motion, but also through extraretinal signals, including efference copy mechanisms and internal forward models that support anticipatory or predictive tracking of moving targets (Spering et al., 2013; Orban de Xivry & Lefèvre, 2007, O’Driscoll & Callahan, 2008; Thakkar & Rolfs, 2019). Smooth pursuit is commonly described as involving an early initiation phase and a later maintenance phase. The initial response is driven primarily by the visual motion signal before feedback from the ongoing eye movement can fully guide performance (open-loop), whereas sustained tracking depends more strongly on feedback, accumulated information about target motion, and internally guided control including predictive signals that support anticipation of the target’s ongoing trajectory (closed-loop). Given the involvement of multiple control signals and response phases, different pursuit measures may capture distinct components of the oculomotor response and reveal different patterns of abnormality.

Impairments in smooth pursuit have long been reported in individuals with psychotic disorders, particularly schizophrenia, and are considered among the most robust and widely replicated oculomotor abnormalities in this population (Holzman et al., 1973, O’Driscoll & Callahan, 2008; Levy et al., 1993). These abnormalities have been characterized using a range of global and specific pursuit measures (Levy et al., 1993; O’Driscoll & Callahan, 2008). Smooth pursuit abnormalities have also been reported in first-degree relatives of individuals with schizophrenia, contributing to the proposal that SPEM deficits may represent an endophenotype of the disorder (Holzman et al., 1974; Thaker et al., 2003; Lencer et al., 2015).

Previous work in schizophrenia has suggested that prediction- or extraretinal driven components of pursuit may be affected (Thaker et al., 1999; Nkam et al., 2010; Spering et al., 2013; Sprenger et al., 2013; Faiola et al., 2020), although findings across specific predictive pursuit measures remain mixed, partly because prediction has been indexed in different ways, through the persistence of eye velocity when target input is removed, or through spatial deviation from the expected target trajectory.

Prediction-related tracking can be evaluated using target disappearance and target blanking paradigms (Lencer et al., 2004; Bennett & Barnes, 2005; Barnes, 2008; Diwakar et al., 2015; Kowler et al., 2019), in which direct visual feedback from the target is temporarily unavailable and continued pursuit must rely more strongly on internal estimates of the target’s ongoing motion. However, relatively few studies have examined pursuit under conditions of structured transient visual occlusion. Paradigms that incorporated target disappearance such as Thaker et al. (1999), typically used blanking procedures in which the target disappeared without a visible occluding object, providing limited spatial structure for evaluating how tracking is maintained during disappearance. Structured occlusion extends this approach by hiding the moving target behind a visible object with clearly defined boundaries, thereby preserving spatial context of the expected target trajectory while temporarily removing direct target information.

A further unresolved issue concerns the sensitivity of smooth pursuit measures to current clinical state. Although several studies have examined oculomotor function across illness stages, most commonly by comparing first-episode with more chronic or remitted presentations (Hutton et al., 1998; Lencer et al., 2010; Lyu et al., 2023), these classifications primarily reflect illness stage rather than the current psychotic state itself. Moreover, such studies have rarely examined whether smooth pursuit abnormalities vary with current psychotic state under conditions of transient visual occlusion. As a result, state-related modulation of smooth pursuit remains insufficiently characterized. Another issue concerns the relationship between smooth pursuit abnormalities and clinical symptom expression. Much smooth pursuit research has been conducted within traditional categorical diagnostic frameworks, most prominently schizophrenia, with relatively limited emphasis on symptom-based or dimensional approaches (Levy et al., 1993; O’Driscoll & Callahan, 2008). As a result, smooth pursuit abnormalities are often described at the level of diagnostic groups rather than in relation to the current clinical expression of psychotic symptoms. Associations between smooth pursuit deficits and the severity of positive or negative symptoms have been examined only to a limited extent, with mixed findings (Ciuffreda et al., 1994; Ross et al., 1996; Kelly et al., 1990; Nkam et al., 2010).

To address these gaps, the current study employed a smooth pursuit paradigm comprising two complementary task contexts: continuous pursuit under uninterrupted visual input and structured transient occlusion, in which tracking had to continue while direct visual feedback from the target was temporarily unavailable. The occlusion condition used a visible occluding object with clearly defined spatial boundaries, a paradigm we have recently applied in Traumatic Brain Injury (Shani et al., in press), allowing assessment of gaze behavior along the expected target trajectory during disappearance. The paradigm was designed to be easy (linear tracking) and divided into short trials, allowing multiple measures of speed, variability, expectation, and inhibition (of eye movements). This design enabled the extraction of pursuit measures not typically captured under conventional visually guided conditions, including occlusion-related deviation, alongside traditional indices of pursuit performance.

Furthermore, we applied this paradigm to individuals across different psychotic clinical states (acute vs. chronic psychosis) and adopted a transdiagnostic approach focused on psychosis as a clinical phenomenon rather than on categorical diagnoses alone. This approach allowed us to examine whether smooth pursuit measures, particularly those obtained during structured occlusion, vary with current psychotic state and symptom profile, and to assess correlations between oculomotor measures and clinical features. Such analyses may reveal state-dependent patterns that complement established trait-oriented accounts of smooth pursuit abnormalities in psychosis.

The present study therefore investigated smooth pursuit in individuals with psychosis using a structured occlusion paradigm, comparing acute and chronic psychotic states with healthy controls and examining whether specific oculomotor measures, including occlusion-related tracking measures, varied with current psychotic state and with positive and negative symptom severity.

## Methods

### Subjects

The study comprised 63 participants: 30 healthy controls (HC) and 33 patients with major psychiatric disorders. Patients were recruited from the psychiatric services of Kaplan Medical Center, including a psychiatric day hospital and outpatient clinics. Healthy controls were recruited from hospital staff and affiliated academic trainees at Kaplan Medical Center. Inclusion criteria were: (1) a diagnosis of a major psychiatric disorder according to DSM-5 criteria (29 schizophrenia-spectrum disorder, 4 bipolar I disorder), based on routine clinical psychiatric assessment by at least two psychiatrists; (2) age 18–75 years; and (3) normal or corrected-to-normal vision sufficient for task performance. Exclusion criteria were: (1) bilateral ocular or oculomotor pathology; (2) extraocular muscle palsy; (3) involuntary head or trunk movements that could interfere with stable positioning in front of the screen and eye tracker; (4) inability to maintain fixation; (5) excessive sleepiness; (6) pregnancy; (7) active substance use; (8) neurological disorders or history of head injury. One patient was excluded during the course of the study due to diagnostic revision (borderline personality disorder) after initially being classified as having a suspected psychotic condition. Patients were classified according to clinical psychotic state into Acute Psychotic (ACPS, n = 16) and Chronic Psychotic (CHPS, n = 17, all with schizophrenia- spectrum disorder) groups.

Acute psychosis was defined as the presence of a current active psychotic episode emerging from a previously non-psychotic or fully remitted baseline, including first-episode psychosis or relapse following a period without psychotic symptoms, typically within the last four weeks. Chronic (residual) psychosis referred to individuals with a continuous baseline expression of psychotic symptoms without full remission.

This classification was defined at the participant level and remained fixed across all study sessions. Symptom severity (PANSS) was assessed at each session and treated as a continuous measure of clinical fluctuation within groups, rather than as a criterion for reclassification. All patients were treated with antipsychotic medications as part of routine clinical care. Medication regimens were determined by the treating psychiatrist and were not experimentally manipulated. Diagnostic breakdown is provided in Table 1.

**Table 1.** Demographic and clinical characteristics of the study groups.

| Group | N | Age<br>(avg ± SD) | Female<br>/ Male | Education<br>(years) | Years of<br>Illness | CGI –<br>Severity | CGI –<br>Improvement | CGI –<br>Efficacy | PANSS –<br>Total | PANSS –<br>Positive | PANSS –<br>Negative |
| --- | --- | --- | --- | --- | --- | --- | --- | --- | --- | --- | --- |
| Control | 30 | 36.73<br>($\pm 12.05$) | 10/20 | 17.21<br>($\pm 2.93$) | N/A | N/A | N/A | N/A | N/A | N/A | N/A |
| Patients All | 33 | 41.09<br>($\pm 14.30$) | 5/28 | 12.42<br>($\pm 1.90$) | 15.72<br>($\pm 10.16$) | 4.92<br>($\pm 0.64$) | 3.13<br>($\pm 0.69$) | 9.98<br>($\pm 2.82$) | 102.19<br>($\pm 20.13$) | 21.45<br>($\pm 7.69$) | 26.59<br>($\pm 7.31$) |
| Acute<br>Psychotic | 16 | 41.94<br>($\pm 17.98$) | 2/14 | 13.00<br>($\pm 2.43$) | 13.36<br>($\pm 11.32$) | 4.88<br>($\pm 0.61$) | 2.83<br>($\pm 0.61$) | 8.53<br>($\pm 2.65$) | 100.48<br>($\pm 20.64$) | 21.63<br>($\pm 7.35$) | 24.84<br>($\pm 7.96$) |
| Chronic<br>Psychotic | 17 | 40.29<br>($\pm 10.23$) | 3/14 | 11.88<br>($\pm 1.02$) | 17.94<br>($\pm 8.68$) | 4.96<br>($\pm 0.69$) | 3.42<br>($\pm 0.64$) | 11.34<br>($\pm 2.28$) | 103.79<br>($\pm 20.13$) | 21.28<br>($\pm 8.21$) | 28.24<br>($\pm 6.44$) |

Values are mean ( ± SD) unless otherwise indicated. “Patients All” comprises the Acute and Chronic psychotic groups combined ( $n = 33$ ); the combined means therefore fall between the two subgroup means.
PANSS subscale values are computed per patient across assessed sessions (un-administered assessments excluded); PANSS–Total = Positive + Negative + General. Female / Male values are counts.
CGI, Clinical Global Impression; PANSS, Positive and Negative Syndrome Scale; SD, standard deviation; N/A, not applicable.

The study was approved by the Kaplan Medical Center Helsinki Committee, and all participants provided written informed consent prior to participation. Patient participants performed the experimental protocol on between one and four separate testing days during their treatment period at Kaplan Medical Center, whereas control participants completed it once. A clinical psychiatric assessment was conducted on each experimental day in the patient group.

### Apparatus

Participants were seated in a stationary chair in front of a laptop display and tested in their hospital room under standard indoor illumination. Head position was not mechanically stabilized (no chin rest), and all recordings were obtained under head-free viewing conditions to allow comfortable bedside testing. Viewing distance was approximately 60 cm but varied across participants and trials. Stimuli were presented on a 14-inch LCD monitor (1366 × 768 pixels, 60 Hz refresh rate) positioned on a standard table. Eye movements and pupil size were recorded binocularly using a remote video-based eye tracker (Tobii Eye Tracker 4C; sampling rate 90 Hz). The Tobii 4C was selected for its robustness and practicality in clinical settings, where rapid setup and tolerance to head movement are essential. Remote video-based eye trackers operating at comparable sampling rates, including Tobii-based systems, have previously been used to extract clinically relevant oculomotor measures—including fixation, pursuit, saccade, vergence, and pupillary measures—in neurological, movement-disorder, and ophthalmologic populations (Cont et al., 2025; Yehezkel et al., 2019; Wygnanski-Jaffe et al., 2023), as well as in our own recent work using the same setup (Shani et al., in press; Meidan & Bonneh, 2026). Although the 90 Hz sampling rate limits fine temporal precision at the single-trial level, it provides sufficient resolution for the spatial and low-frequency dynamics examined here. In particular, temporally sensitive measures (e.g., initial catch-up saccade latency) were interpreted as trial-averaged estimates, which are coarse per trial but become reliable at the participant level through repeated measurements. Many key outcome measures—such as tracking gain, deviation during occlusion, and pupil dynamics—primarily depend on slower or spatially defined signals and are therefore less sensitive to sampling rate limitations. Stimuli were generated using an in-house platform for psychophysical and eye-tracking experiments (PSY), developed in our laboratory and used in previous studies (e.g., Bonneh et al., 2015; Kadosh & Bonneh, 2022; Ziv & Bonneh, 2021).

### Stimuli and procedure

#### Stimuli

Two smooth-pursuit paradigms were used (Figure 1): a standard visible-target condition (Experiment 1) and an occluded-target condition (Experiment 2). Each trial began with a central fixation cross (∼1° visual angle) presented for 500 ms on a black background. A bright circular target (∼1.5° diameter) then appeared at fixation and moved smoothly outward for 1.5 s along a straight trajectory. Motion direction was selected randomly on each trial from eight possible directions (four cardinal and four diagonal). Target motion was defined in screen coordinates such that the horizontal and vertical velocity components were constant (∼5.7°/s at a nominal viewing distance of 60 cm). As a result, diagonal motion had a higher resultant speed (∼8.1°/s). This design ensured identical component velocities across directions while preserving isotropy of motion trajectories. The relatively large target size was chosen to ensure robust visibility and stable tracking under head-free bedside conditions, rather than to probe fine spatial limits of pursuit (Heinen & Watamaniuk, 1998). Each run included 16 trials (two per direction) with a 500 ms inter-trial interval and lasted approximately 0.75 minutes.

**Figure 1.**
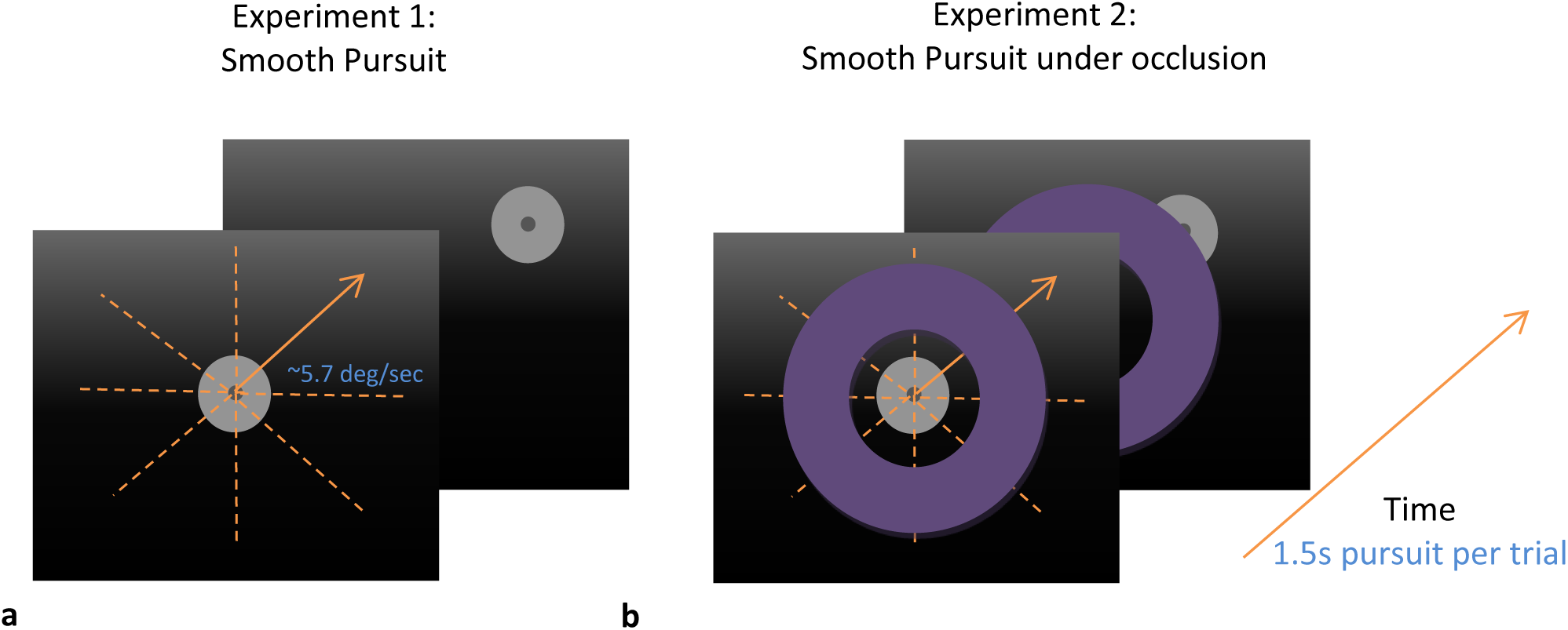
Stimuli used for the smooth pursuit experiments. (**a**) Simple pursuit. A central fixation cross was shown for 500 ms, after which a bright disk appeared at fixation and moved smoothly outward for 1.5 s along one of eight randomly selected directions. Target motion was defined by horizontal and vertical velocity components of ∼5.7°/s at a nominal viewing distance of 60 cm. Each run included 16 trials. (**b**) Occluded pursuit. The same sequence was used, with the addition of an occluding ring centered on fixation. The occluder had one of three widths; the medium-width occluder is shown.

The occlusion paradigm (Experiment 2) was identical in structure to the standard pursuit condition, except that the moving target passed behind a circular occluding region centered on fixation. The occluder was a bright magenta annular region (luminance ∼21.6 cd/m², Figure 1b) with three possible widths (∼2°, ∼4°, and ∼6°), presented in random order (one trial per direction for each width; 24 trials per run, ∼1.6 min).

#### Procedure

Participants were instructed to follow the moving target as accurately as possible with their eyes. No additional task or response was required. A standard multi-point calibration procedure (Tobii) was performed at the beginning of each session. Participants were seated at approximately 60 cm from the display, although this distance varied due to the head-free setup and clinical constraints. Each unoccluded run lasted approximately 0.75 minutes and each occluded run approximately 1.6 minutes; each run type was repeated three times, for a total recording time of about 7 minutes per session. Patient participants completed between one and four sessions on separate days during their treatment period, whereas control participants completed a single session. Clinical assessments (PANSS) were obtained on each testing day and used to characterize symptom severity.

### Data Analysis

#### Preprocessing

Because recordings were obtained under head-free viewing conditions, the viewing distance varied across participants and trials. For each trial, viewing distance was estimated from the eye tracker signal during the pre-stimulus interval (−0.5 to 0 s), excluding outliers (>2 SD). Trials with extreme distances (<40 cm or >80 cm) were discarded. All gaze data were converted from screen coordinates to degrees of visual angle using a nominal viewing distance of 60 cm, allowing comparison across participants. This approximation introduces only minor scaling variability and does not affect the relative pattern of results. Periods of missing data due to blinks or tracking loss were identified based on loss of pupil signal and confirmed using vertical gaze deviations. These segments were removed from further analysis. The proportion of valid samples was analyzed across groups as an index of recording quality (see Results).

#### Saccade Detection

Saccades were detected using a velocity-threshold method based on the framework of Engbert and Kliegl (2003), which applies an adaptive, noise-dependent criterion. Although originally developed for microsaccades, this approach is widely used for detecting saccadic events across a broad amplitude range when parameters are appropriately adjusted. Given the sampling rate (90 Hz) and noise characteristics of the Tobii 4C, velocity thresholds of 8–400°/s were applied to exclude noise and non-physiological events while retaining tracking-related catch-up saccades. No lower amplitude threshold was imposed, as the primary analyses were based on trial-averaged measures that are robust to occasional false detections. We also did not impose the conventional lower peak-velocity cutoff (e.g., 30°/s), because moderate sampling rates can attenuate estimated peak velocities for small saccades.

To exclude large reorienting movements, an upper amplitude limit of 8° was applied. Detected events exhibited the expected main-sequence relationship between amplitude and peak velocity, supporting their physiological validity. Importantly, the pattern of group differences was stable across a range of detection parameters.

#### Smooth and Saccadic Pursuit

To separate smooth and saccadic components of tracking, saccades were removed from the gaze traces by marking saccade periods as missing and aligning the surrounding segments by subtracting the saccade-induced displacement. The resulting traces represent the smooth pursuit component. The saccadic component (“saccadic pursuit”) was defined as the difference between the full tracking trace and the saccade-free trace. Pursuit speed for both components was estimated using robust linear fits within predefined time windows, and expressed as gain relative to target speed.

#### Oculomotor Measures

Several oculomotor measures were derived from the tracking data, computed per trial and then averaged within participant after outlier rejection (>2 SD, applied twice):

- <u>Smooth tracking gain</u>: Estimated from the velocity of the saccade-free trace relative to target velocity (typically 0.5–1 s after motion onset; Figures 2–3).
- <u>Tracking Deviation</u>: Radial deviation of gaze position relative to target trajectory during sustained tracking.
- <u>Pupil response</u>: Percent change in pupil size relative to pre-trial baseline, quantified over predefined time windows.
- <u>Occluder-Induced Deviation</u>: To combine motion directions without sign cancellation, horizontal and vertical gaze traces were first baseline-corrected at motion onset (t = 0) and transformed to unsigned values. For each occluded trial, deviation from the target position time-course reference was computed and averaged over two predefined windows, 0.5–1.0 s and 1.0–1.5 s after motion onset. The resulting deviation measure was computed as the radial gaze distance minus the radial target distance, so that positive values indicate a predictive lead relative to the expected target trajectory and negative values indicate a lag.

**Figure 2.**
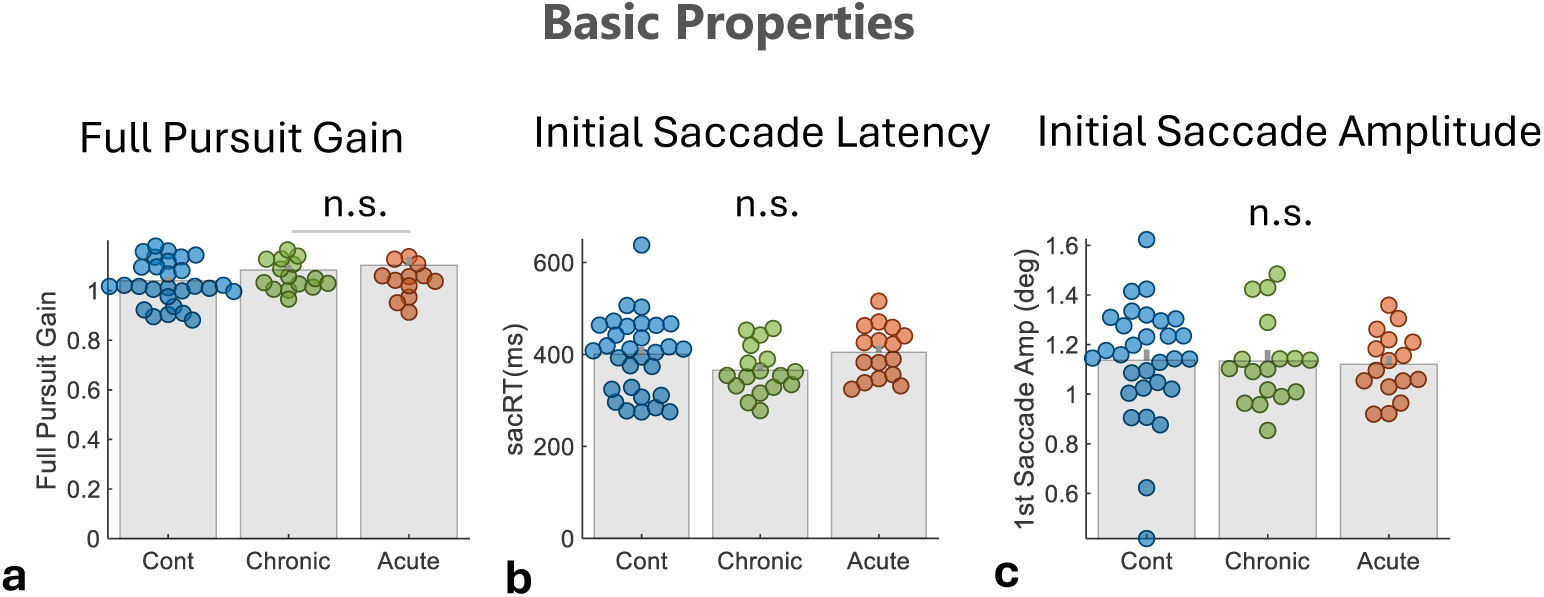
Basic oculomotor measures of unoccluded pursuit in patients and controls. (a) Full pursuit gain, computed as the ratio of pursuit velocity, including catch-up saccades, to target velocity, assessed in the 0.5–1.0 s window after target-motion onset in Experiment 1. (b) Initial catch-up saccade latency relative to target-motion onset. (c) Initial catch-up saccade amplitude. The initial catch-up saccade was identified in the 0.25–0.75 s window after target-motion onset. Each dot represents the participant mean across trials after exclusion of outlier measures (>2 SD; see Methods), and bars denote group means. Group differences were minimal: initial catch-up saccade latency and amplitude did not differ between groups, whereas full pursuit gain showed a weak, borderline group effect, with slightly higher values in acute patients than in controls — opposite in direction to a deficit (see Results).

**Figure 3.**
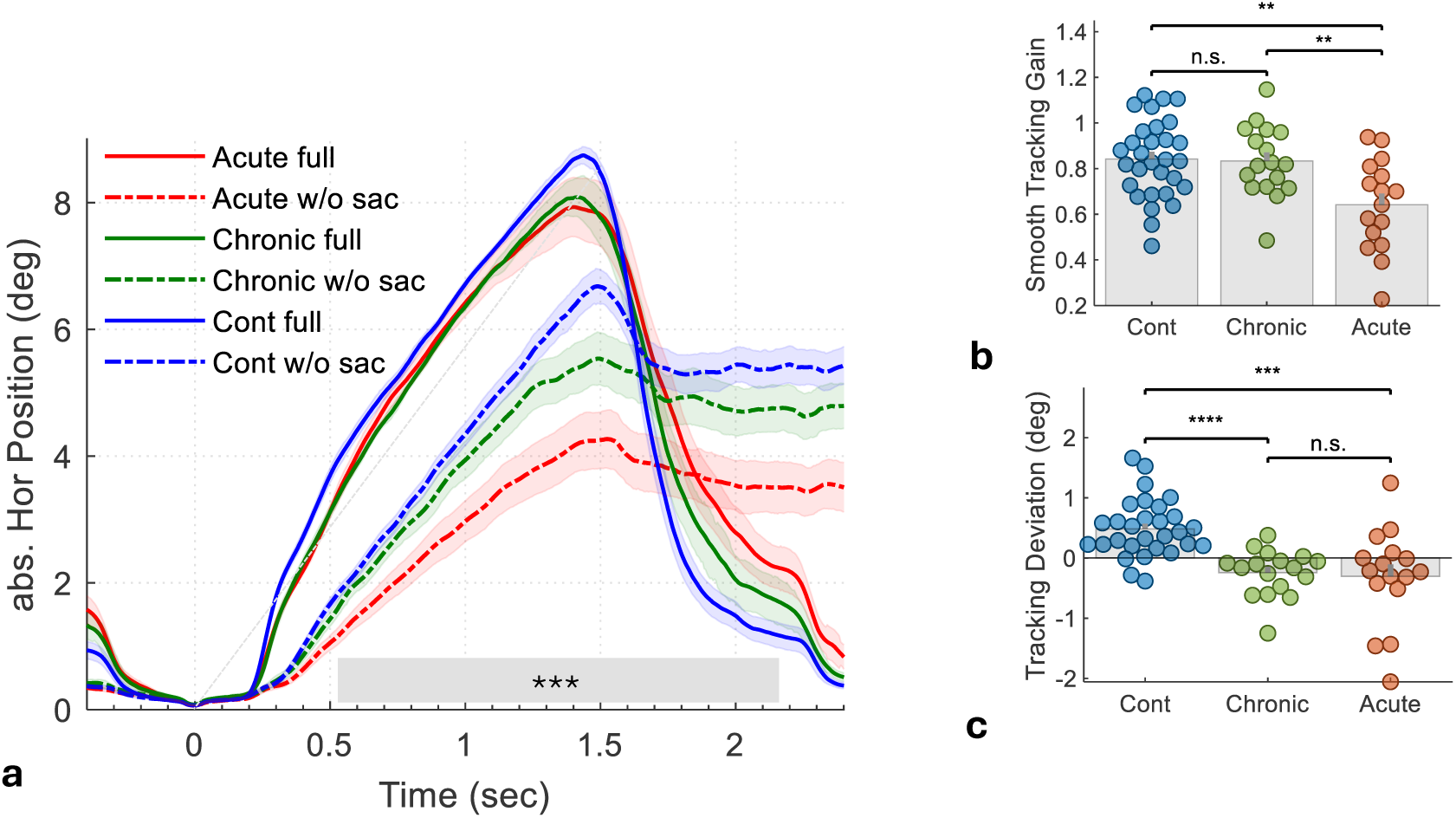
Results for smooth and catch-up saccade pursuit (Experiment 1). **(a)** Horizontal gaze traces as a function of time, normalized by time 0 and converted to absolute values, averaged across trials of all directions excluding vertical (with outlier rejection) and then across observers, for the full traces (solid lines) and for traces without saccades (after saccade removal and alignment, dashed lines), for the three groups. Error bars denote ±1 SE across observers. The smooth (without saccades) part of tracking (dashed lines) was compared between the Cont (blue) and Acute (red) groups via a permutation test, yielding p < .001 (gray bar). The light blue line shows the stimulus position over time. **(b)** Bee-swarm plot of smooth tracking gain (estimated speed of the saccade-free traces relative to stimulus speed in experiment 1, in 0.5-1s post target onset). Group differences were evaluated using a linear mixed-effects model on trial-level data with random intercepts for participant and session nested within participant. The model revealed a significant main effect of Group. Acute patients showed significantly reduced gain relative to both Controls and Chronic patients, whereas Chronic patients did not differ from Controls. **(c)** Bee-swarm plot of tracking deviation (radial, see Methods). A linear mixed-effects model revealed a significant main effect of Group. Both Chronic and Acute patients showed a significantly negative deviation (small lag) relative to Controls who were slightly ahead of the target, whereas the two patient groups did not differ from each other.

All measures were computed per condition and then averaged across sessions to yield a single value per participant for group-level analyses, unless otherwise specified.

#### Time-course Measures

Time-course analyses were performed for gaze position, pupil size, and saccade rate. Saccade-rate modulation functions were computed by convolving saccade onset events with a Gaussian kernel (σ = 50 ms), yielding a continuous estimate of event rate over time. Eye velocity time courses were computed using sliding windows (150 ms width, 22 ms step), based on two-dimensional displacement. To allow averaging across directions, diagonal velocities were normalized by √2 to match cardinal component speeds.

### Statistical Analysis

Statistical analyses were performed in MATLAB (R2024b). Unless otherwise stated, significance was defined as α = 0.05 (two-sided). Group differences in oculomotor measures were assessed using linear mixed-effects (LME) models applied to trial-level data, with group as a fixed effect and random intercepts for participant and session nested within participant. Time-resolved comparisons were evaluated using cluster-based nonparametric permutation tests, controlling multiple comparisons across time. Associations with symptom severity (PANSS) were analyzed at both the participant level (using Pearson correlation) and the observation level (patient-by-session) using LME models. For visualization, orthogonal regression was used to account for variability in both variables, with confidence intervals estimated via bootstrap resampling (1,000 iterations).

## Results

Participants performed smooth pursuit tracking of a moving target (Experiment 1) and tracking under occlusion (Experiment 2; Figure 1). In the following, we report the results for the different oculomotor measures and then their correlation with the clinical symptoms.

### Eye-tracking data quality

The percentage of valid gaze samples in the full recordings, excluding blinks and off-screen artifacts, was high across the cohort (M = 81%, SD = 13.5). A linear mixed-effects model showed no significant omnibus effect of Group, F(2,642) = 2.75, p = .06. Controls had a slightly higher proportion of valid samples than Acute patients, β = 0.093, SE = 0.040, t(642) = 2.32, p = .02, whereas Chronic patients did not differ from either group, ps ≥ .23. Thus, recording quality was broadly comparable across groups, with a small advantage for Controls relative to Acute patients.

To quantify tracker precision under the head-free recording conditions, we computed the root-mean-square sample-to-sample displacement of gaze position during the pre-target fixation baseline period, −0.5 to 0 s relative to target-motion onset, after removal of blinks, missing samples, and saccadic segments (RMS S2S). Mean RMS S2S values after excluding two extreme outlier participants (>4 SD; one Chronic patient and one Control) were low across groups: 0.065 ± 0.025° in Acute patients, 0.082 ± 0.022° in Chronic patients, and 0.067 ± 0.023° in Controls. A linear mixed-effects model applied to trial-level RMS S2S values showed no significant omnibus group effect, F(2,12840) = 2.53, p = .08. Chronic patients showed higher RMS S2S than Controls, β = 0.024, SE = 0.012, t ≈ 2.06, p = .04, and a trend toward higher RMS S2S than Acute patients, β = 0.025, SE = 0.013, t(12840) = 1.90, p = .06. Acute patients and Controls did not differ, p = .93. Thus, tracker precision was generally good under the head-free bedside conditions, and the main Acute-patient effects are unlikely to be explained by poorer gaze precision in the Acute group.

### Basic oculomotor measures

We first examined basic oculomotor measures that reflect task engagement and overall tracking performance (Figure 2). Full pursuit gain showed only a weak group effect in a linear mixed-effects model on trial-level data, F(2,4229) = 2.96, p = .05, driven by slightly lower values in controls than in acute patients (β = −0.061, SE = 0.026, t(4229) = −2.37, p = .02); chronic patients did not differ from either acute patients (p = .42) or controls (p = .14). Neither initial catch-up saccade latency (F(2,2337) = 1.50, p = .22) nor initial catch-up saccade amplitude (F(2,2337) = 0.24, p = .79) showed a significant effect of Group. Thus, basic oculomotor measures were largely comparable across groups, with no evidence for a generalized deficit in task engagement, pursuit initiation, or the initial catch-up response. Although single-trial saccade timing is measured with limited precision at 90 Hz, this latency measure is dominated by the first directionally appropriate catch-up saccade following motion onset and is therefore robust at the participant level to occasional early or small-amplitude detections.

### Smooth tracking gain

The results for smooth tracking gain are shown in Figure 3. As illustrated in Figure 3a, the overall tracking trajectories (solid lines) were broadly similar across groups, whereas the smooth pursuit component obtained after removing saccades and realigning the remaining trajectory (dashed lines) differed markedly between groups. To account for repeated trials and repeated testing sessions when assessing smooth tracking gain (Figure 3b), we fitted a linear mixed-effects model to trial-level gain values (N = 3,611), with Group as a fixed effect and random intercepts for participant and session nested within participant. The model revealed a significant main effect of Group (F(2,3608) = 5.995, p = .003). Relative to Controls, Acute patients exhibited significantly reduced gain (β = −0.157, SE = 0.050, t(3608) = −3.14, p = .002), whereas Chronic patients did not differ from Controls (β = 0.003, SE = 0.050, t(3608) = 0.07, p = .95). Direct comparison between the two patient groups confirmed lower gain in Acute relative to Chronic patients (β = −0.161, SE = 0.055, t ≈ −2.94, p = .003). Together, these results indicate a selective reduction in smooth tracking gain during the acute phase of illness, with chronic patients showing gain values comparable to those of controls.

### Tracking Deviation

The results for the tracking deviation measure are shown in Figure 3c. Tracking deviation was analyzed during Experiment 1 (unoccluded tracking) using a linear mixed-effects model applied to trial-level data (N = 4,539), with Group as a fixed effect and random intercepts for participant and session nested within participant. The model revealed a significant main effect of Group (F(2,4536) = 17.00, p < .0001). Relative to acute patients, controls showed significantly higher tracking deviation values (β = 0.811, SE = 0.171, t(4536) = 4.74, p < .0001), reflecting a more predictive tracking pattern with gaze positioned further ahead of the moving target. Chronic and acute patients did not differ from one another (β = −0.040, SE = 0.189, t(4536) = −0.21, p = .83). Controls also differed significantly from chronic patients (β = 0.851, SE = 0.171, t ≈ 4.98, p < .0001). Together, these results indicate that both acute and chronic patients exhibited a reduced predictive lead relative to controls, while the two patient groups showed comparable levels of impairment. This pattern suggests that altered predictive tracking may reflect a broader psychosis-related alteration, present across acute and chronic states, rather than a purely state-dependent effect.

### Tracking under occlusion

The results for occluded tracking (Experiment 2) are shown in Figure 4. The effect of target occlusion can be appreciated in the horizontal tracking traces (all directions except vertical; Figure 4a). During the occlusion period, control participants continued to track predictively and moved ahead toward the anticipated target reappearance location, whereas both patient groups showed a reduced anticipatory response. This difference was most pronounced in the acute patient group, whose gaze remained substantially behind the expected target position after target reappearance. To quantify this effect, we computed radial tracking deviation relative to the target trajectory (Figure 4b, widest occluder). Positive values indicate a predictive lead relative to the target, whereas negative values indicate lag. The time course demonstrates a clear separation between groups during the occlusion period, with controls exhibiting the largest predictive lead, chronic patients showing an intermediate response, and acute patients exhibiting the greatest lag.

**Figure 4.**
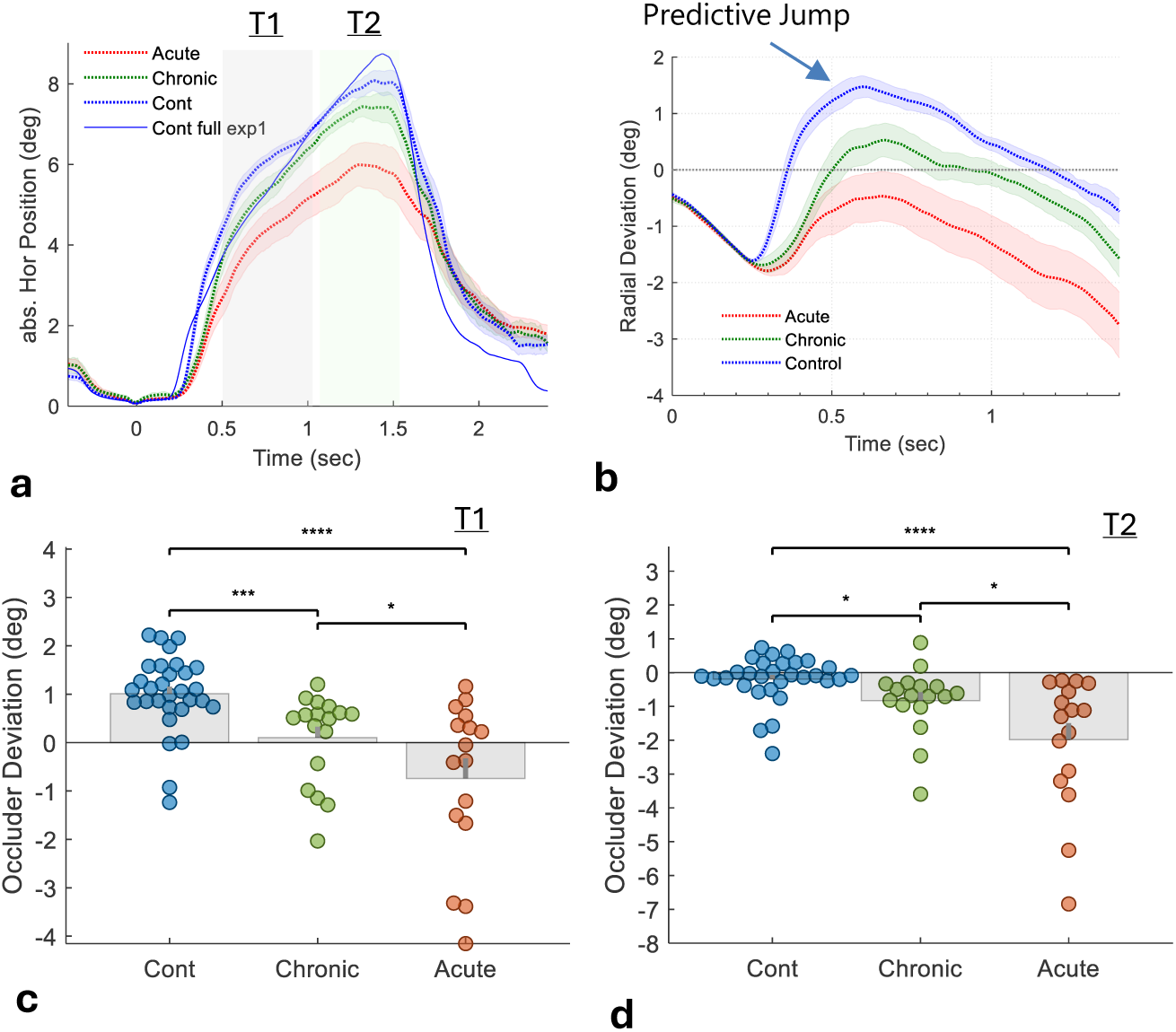
Smooth pursuit under occlusion (Experiment 2). (a) Horizontal gaze traces (all directions except vertical) as a function of time, normalized to time 0 and expressed in absolute position, averaged across trials and then across observers (±1 SE). Curves show Acute (red), Chronic (green), and Control (blue) groups during occluded tracking with the large occluder. The solid blue curve shows the Control group average from unoccluded tracking in Experiment 1. (b) Radial deviation from the target movement trajectory, computed as the difference in radial gaze distance (√(H²+V²)). Negative values indicate lag relative to the target trajectory. Curves show group means ±1 SE across observers. (c–d) Group comparison of mean occluder-induced radial deviation for the large occluder, computed in the early window, 0.5–1.0 s, and late window, 1.0–1.5 s, respectively. Individual dots represent observers and bars denote group means. Statistical comparisons were performed using linear mixed-effects models on trial-level data with random intercepts for participant and session nested within participant. See Results for the LME results.

We quantified these differences in two temporal intervals during occlusion: an early window (0.5–1.0 s; T1) and a late window (1.0–1.5 s; T2), using linear mixed-effects models applied to trial-level data, with Group as a fixed effect and random intercepts for participant and session nested within participant (Figure 4c–d). In the early window (Figure 4c; N = 2,137), the model revealed a significant main effect of Group (F(2,2134) = 14.14, p < .0001). Relative to acute patients, chronic patients (β = 0.81, SE = 0.39, t(2134) = 2.07, p = .04) and controls (β = 1.82, SE = 0.35, t(2134) = 5.21, p < .0001) showed significantly higher tracking deviation values. Controls also differed significantly from chronic patients (β = 1.01, SE = 0.35, t ≈ 2.93, p = .003).

A similar pattern was observed in the late window (Figure 4d; N = 1,993), with a significant main effect of Group (F(2,1990) = 11.84, p < .0001). Relative to acute patients, both chronic patients (β = 0.89, SE = 0.40, t(1990) = 2.20, p = .03) and controls (β = 1.76, SE = 0.36, t(1990) = 4.83, p < .0001) showed significantly higher values. Controls also differed significantly from chronic patients (β = 0.87, SE = 0.36, t ≈ 2.41, p = .02). Together, these results indicate a graded impairment in predictive tracking during target occlusion, with controls showing the strongest anticipatory tracking, chronic patients an intermediate level of performance, and acute patients the weakest predictive response.

### Pupillary responses

We next examined pupillary responses during occluded tracking as an index of task-evoked autonomic and visual responsivity (Figure 5). We focused on Experiment 2 because the bright occluder elicited a robust pupil light response, providing a clear condition for comparing pupil constriction across groups. As shown in Figure 5a–c, controls exhibited stronger pupil constriction during and following target motion, whereas both patient groups showed reduced constriction. Time-course analysis revealed significant group differences during the late portion of the trial (cluster-based permutation test, p < .001), primarily reflecting differences between controls and acute patients. To quantify these effects, we computed the late pupil response as the mean percent change in pupil size relative to the pre-trial baseline during the 1.5–2.0 s interval and analyzed this trial-level measure using linear mixed-effects models with Group as a fixed effect and random intercepts for participant and session nested within participant.

**Figure 5.**
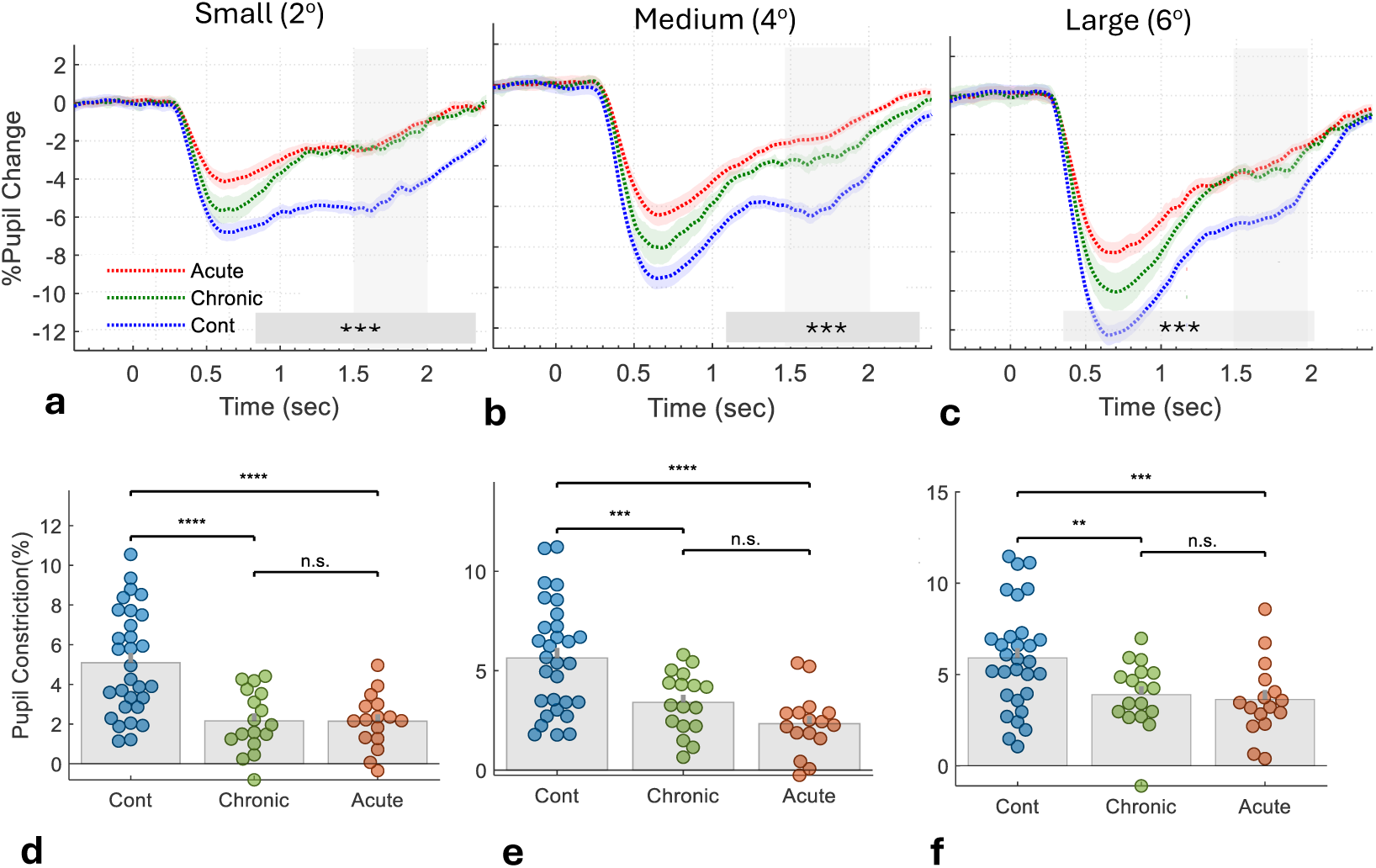
Pupillary responses during occluded smooth pursuit. (a–c) Time course of percent change in pupil size relative to the pre-trial baseline for the three groups in Experiment 2, shown separately for the three occluder widths: small (∼2°), medium (∼4°), and large (∼6°). Curves show group means ±1 SE across observers for Controls (blue), Chronic patients (green), and Acute patients (red). Shaded gray intervals indicate time periods with significant group differences (cluster-based permutation test, p < .001; dominant contrast: Controls vs. Acute). (d–f) Group comparison of late pupil response, quantified as the average percent change relative to baseline in the 1.5–2.0 s post-stimulus interval indicated by the gray windows in panels a–c. For clarity, constriction values are plotted as positive. Panels d–f correspond to panels a–c, respectively. Individual points represent observers and bars denote group means.

For the small occluder condition (Figure 5d; ∼2°), the model revealed a significant main effect of Group, F(2,2187) = 20.54, p < .0001. Relative to Acute patients, Controls showed stronger constriction, β = −3.116, SE = 0.568, t(2187) = −5.49, p < .0001, whereas Chronic patients did not differ from Acute patients, β = −0.138, SE = 0.622, t(2187) = −0.22, p = .82. Controls also showed stronger constriction than Chronic patients, β = −2.978, SE = 0.569, t ≈ −5.24, p < .0001. A similar pattern was observed for the medium occluder condition (Figure 5e; ∼4°), with a significant main effect of Group, F(2,2217) = 12.41, p < .0001. Controls showed stronger constriction than Acute patients, β = −3.031, SE = 0.631, t(2217) = −4.81, p < .0001, and stronger constriction than Chronic patients, β = −2.026, SE = 0.636, t ≈ −3.19, p = .001. Chronic and Acute patients did not differ significantly, β = −1.006, SE = 0.686, t(2217) = −1.47, p = .14. For the large occluder condition (Figure 5f; ∼6°), the Group effect also remained significant, F(2,2217) = 7.35, p < .001. Controls again showed stronger constriction than Acute patients, β = −2.557, SE = 0.714, t(2217) = −3.58, p < .001, and stronger constriction than Chronic patients, β = −1.888, SE = 0.711, t ≈ −2.66, p = .008. Chronic and Acute patients did not differ significantly, β = −0.670, SE = 0.788, t(2217) = −0.85, p = .40.

Together, these results indicate that pupil constriction in occluded tracking was reduced in both patient groups relative to controls across occluder widths, with no reliable difference between Acute and Chronic patients.

### Correlation with PANSS symptoms

To examine the relationship between oculomotor performance and clinical symptom severity, we correlated each measure with PANSS scores (Figure 6). The figure presents both patient means across testing days (left column) and all patient-by-session observations (right column). Statistical significance for the repeated-measures data was assessed using linear mixed-effects (LME) models with random intercepts for participant.

**Figure 6.**
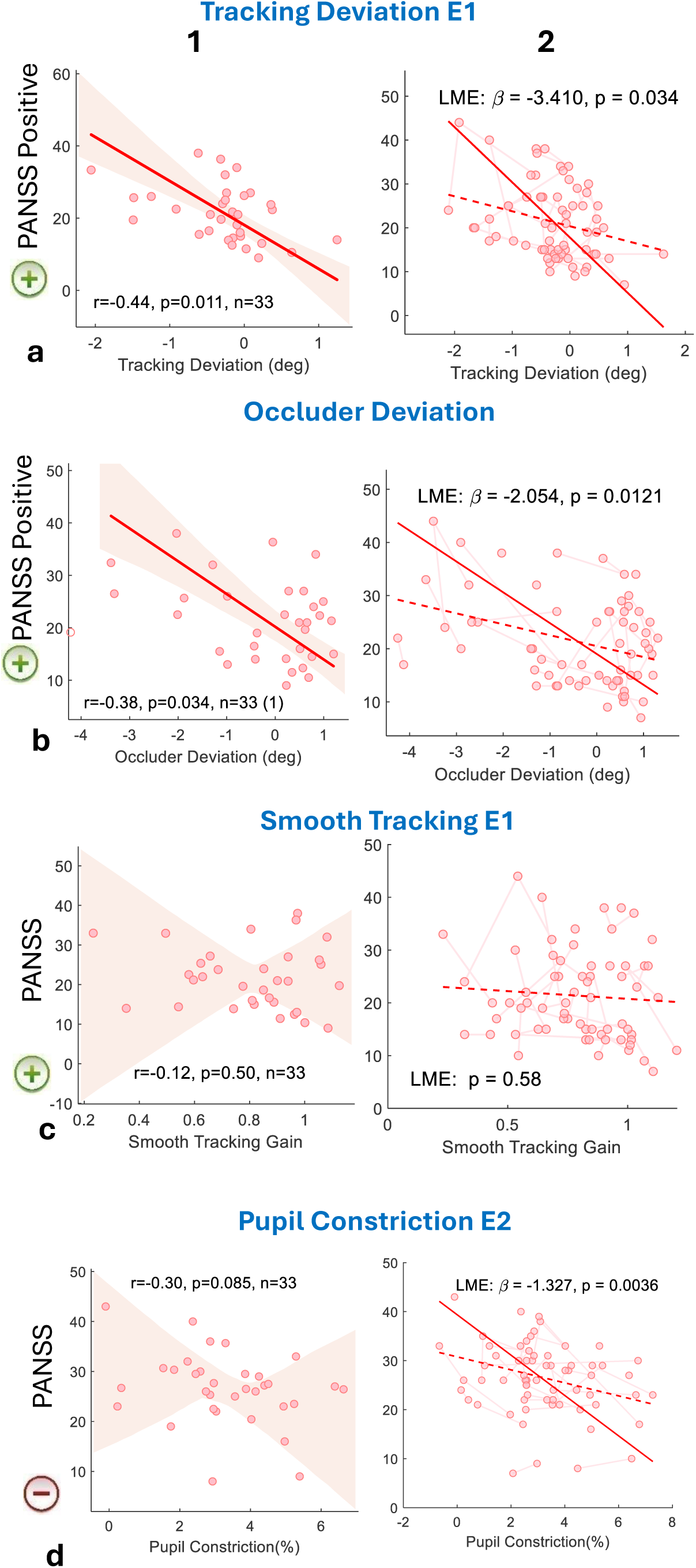
Oculomotor measures and symptom severity. Four oculomotor measures are shown: (a) tracking deviation during unoccluded tracking (Experiment 1), (b) occluder deviation during predictive tracking under occlusion (Experiment 2); Positive values indicate a predictive lead relative to the target trajectory, whereas negative values indicate lag behind the target. (c) smooth tracking gain during unoccluded tracking (Experiment 1), and (d) pupil constriction during occluded tracking (Experiment 2). Pupil constriction was quantified as the mean late pupil response in the 1.5–2.0 s interval of Experiment 2, averaged across the medium and large occluder-width conditions. For each measure, associations with PANSS symptom severity are shown in two formats: patient means across testing days (left column; one point per patient) and patient-by-session observations (right column). Observations from the same patient are connected by light pink lines. Solid red lines indicate orthogonal-regression fits (left column) or linear mixed-effects model fits (right column). Dashed red lines indicate ordinary least-squares fits for visualization. Shaded regions show 95% bootstrap confidence intervals around the orthogonal-regression fits. Pearson correlation coefficients (r) are shown for patient means, and linear mixed-effects model coefficients (β) and p-values are shown for the patient-by-session analyses.

Measures reflecting predictive-tracking performance showed significant negative associations with PANSS positive symptoms. Tracking deviation during unoccluded tracking (Experiment 1) was negatively correlated with positive symptoms at the patient-mean level (*r* = −0.44, *p* = .011, *n* = 33). The corresponding patient-by-session analysis confirmed this relationship (F(1,64) = 4.72, *p* = .03; β = −3.41, SE = 1.57, *t*(64) = −2.17; *N* = 66 observations). Similarly, occluder deviation during predictive tracking under occlusion (Experiment 2) was negatively associated with PANSS positive symptoms both at the patient-mean level (*r* = −0.38, *p* = .034, *n* = 33) and in the patient-by-session analysis (F(1,63) = 6.67, *p* = .01; β = −2.05, SE = 0.80, *t*(63) = −2.58; *N* = 65 observations). Because higher deviation values reflect a larger predictive lead relative to the target trajectory, these findings indicate that more severe positive symptoms were associated with reduced predictive tracking and greater lag relative to the target. In contrast, smooth tracking gain during unoccluded tracking (Experiment 1), a measure commonly reported in schizophrenia research, was not associated with PANSS positive symptoms either at the patient-mean level (*r* = −0.12, *p* = .50, *n* = 33) or in the patient-by-session analysis (F(1,64) = 0.31, *p* = .58; β = −2.92, SE = 5.29, *t*(64) = −0.55; *N* = 66 observations).

Pupil constriction in occluded tracking assessed in the post-stimulus period, quantified as the mean late pupil response (1.5–2.0 s) averaged across the medium and large occluder-width conditions, was negatively associated with PANSS negative symptoms. At the patient-mean level, the association was in the expected direction but did not reach conventional significance (*r* = −0.30, *p* = .085, *n* = 33). However, the patient-by-session analysis revealed a significant negative relationship (F(1,63) = 9.14, *p* = .004; β = −1.33, SE = 0.44, *t*(63) = −3.02; *N* = 65 observations), indicating that stronger pupil constriction measured post-stimulus presentation was associated with lower negative symptom severity.

Together, these findings suggest that symptom severity is more closely related to measures of predictive tracking and pupil response than to smooth pursuit gain. More severe positive symptoms were associated with reduced predictive tracking, whereas more severe negative symptoms were associated with weaker pupil constriction.

## Discussion

### Overview of findings

The present study examined smooth pursuit in patients with acute and chronic psychosis using continuous tracking and structured target occlusion. During continuous pursuit, the control group showed a slight forward tracking pattern, whereas both patient groups showed a small lag relative to controls. This group difference occurred despite largely preserved other pursuit-related measures, suggesting a specific difference in tracking pattern with the moving target rather than a generalized difference in pursuit performance. The structured occlusion condition extended this finding by showing reduced forward progression of gaze during target disappearance and greater positional lag after reappearance. Importantly, abnormalities were not uniform across measures or clinical states: tracking deviation effects were present in both

Acute and Chronic psychosis, whereas smooth tracking gain and occluder deviation during both target disappearance and post-reappearance tracking were more pronounced during acute psychosis. The trajectory-based measures were also associated with current positive symptom severity, supporting their clinical relevance. Together, these findings suggest a broader psychosis-related alteration in spatial tracking, accompanied by additional state-sensitive disruptions in specific components of pursuit performance.

### Preserved basic pursuit with altered sustained tracking and gaze–target alignment

Basic pursuit-related measures were largely preserved across groups. During unoccluded pursuit, the latency and amplitude of initial catch-up saccades did not differ between groups, and full pursuit gain showed only a weak, borderline group effect, slightly higher in Acute patients than in controls (Figure 2). Because full pursuit gain includes catch-up saccades, this small difference runs opposite to what a deficit would produce and most plausibly reflects preserved or compensatory saccadic catch-up rather than superior smooth pursuit; it also contrasts with the reduced saccade-free smooth gain in Acute patients described below. The group differences seen in our later analyses are therefore unlikely to reflect a generalized oculomotor deficit or a broad inability to perform the task. The differences that did emerge were instead confined to more specific measures of sustained tracking and gaze–target alignment. Smooth tracking gain, computed from the saccade-free traces, was selectively reduced in the Acute group, whereas the Chronic group did not differ from controls (Figure 3), suggesting that smooth velocity matching may be particularly vulnerable during acute psychotic states. Tracking deviation, by contrast, was altered in both Acute and Chronic patients, including the Chronic group, whose smooth tracking gain was preserved. It therefore cannot be explained by reduced smooth gain alone, and more likely reflects a broader difference in how gaze advances along the target trajectory, with patients showing reduced forward displacement relative to the control tracking pattern.

### Occlusion-related tracking and post-occlusion realignment

Structured occlusion further revealed altered tracking when direct visual feedback from the target was temporarily unavailable. During occlusion, control participants showed a predictive jump (jump ahead) relative to the unoccluded reference trajectory, consistent with tracking along the expected target path. In contrast, both patient groups showed reduced forward progression, with Acute patients showing the strongest reduction and Chronic patients showing an intermediate response. This suggests that the ability to maintain gaze along the expected path during temporary target disappearance was modulated by current clinical state. (Figure 4a–b) A similar state-sensitive pattern was observed after target reappearance: Acute patients showed the largest positional lag, whereas Chronic patients showed an intermediate profile. Thus, the occlusion paradigm extended the continuous-pursuit findings by showing how gaze–target alignment is maintained and re-established when visual feedback from the target is interrupted.

### Prediction-related interpretation of the results

An important question is whether the current findings reflect altered prediction-related tracking. Smooth pursuit integrates visual feedback with extraretinal and internally generated signals that compensate for visuomotor delays and support anticipatory tracking (Barnes, 2008). Predictive pursuit includes both eye movements toward expected future target locations and continued tracking when direct target information is temporarily unavailable (Kowler et al., 2019). Moreover, eye velocity can recover before an occluded target reappears, demonstrating predictive extrapolation during target disappearance (Bennett & Barnes, 2005). In the present study, tracking showed a graded reduction in forward progression under occlusion, with the largest reduction in Acute patients (Figure 4a–d), consistent with impaired maintenance or updating of the expected target trajectory. The group traces indicate how this pattern arises: during occlusion, control participants generated a discrete, spatially targeted movement toward the occluder exit and then waited, producing a forward predictive lead that is visible approximately 0.5–1 s after motion onset in the occluded tracking traces (Figure 4a). In contrast, patients did not show this anticipatory “jump ahead and wait” response. The chronic group, in which smooth gain was relatively preserved, instead continued to track with a more maintained velocity. This same difference may help reconcile our findings with prior work that at first appears to point in the opposite direction: using target blanking, Sprenger et al. (2013) reported slower eye deceleration after target extinction in schizophrenia, interpreted as greater—rather than reduced—reliance on predictive signals. The two accounts are compatible once the measures are distinguished, because Sprenger et al. indexed velocity persistence when visual input was removed, whereas our measure indexes spatial deviation relative to the expected trajectory in the presence of a visible occluder with a defined reappearance location. A continuation of ongoing eye velocity in the absence of the target is precisely the pattern that yields the slower deceleration they reported. One possibility is that controls deploy a discrete, spatially precise prediction (moving ahead and waiting), whereas patients rely more on lower-level velocity extrapolation, which would appear as enhanced persistence in a velocity metric but as a reduced predictive lead in a position metric. The present positional traces are consistent with this account but do not by themselves establish it. In the Acute group, the larger positional lag co-occurred with reduced smooth gain (Figure 3b) and thus likely reflects an additional velocity deficit rather than persistence alone.

It should be noted that the current paradigm does not isolate prediction per se, because performance under occlusion may also depend on attention, sensorimotor updating, short-term motion memory, and arousal. The findings therefore support altered prediction-related tracking rather than demonstrating a pure predictive deficit, consistent with previous empirical and theoretical work in schizophrenia (Nkam et al., 2010; Adams et al., 2012; Sprenger et al., 2013).

The graded group effect in the occlusion experiment, together with the association of both tracking deviation and occlusion-related deviation with positive symptom severity, suggests that these measures may warrant investigation as state-sensitive markers, potentially indexing dynamic changes in psychotic-state expression rather than stable trait abnormalities alone. Repeated assessments could potentially help monitor clinical improvement or treatment response. However, given the modest sample size and the absence of a longitudinal treatment design, further longitudinal work is needed to evaluate this possibility.

### Clinical-state and symptom-related effects

Beyond the group differences summarized above, the trajectory-based measures were related to current symptom severity. Both continuous tracking deviation and occluder deviation were associated with positive symptom severity in patient-mean and patient-by-session analyses (Figure 6a–b), whereas smooth tracking gain was unrelated to positive symptoms (Figure 6c). These associations suggest that positive symptoms are more closely linked to trajectory-based measures of forward and predictive tracking than to conventional smooth pursuit gain.

### Pupillary responses

The pupillary findings reported here are secondary and exploratory; the present task was not designed as a dedicated pupillometry paradigm, and these results are offered as a complementary physiological observation. In addition to the smooth pursuit findings, patients showed reduced pupil constriction relative to controls during occluded tracking across all three occluder-width conditions (Figure 5). Pupil constriction varied with occluder size, with larger occluders eliciting stronger constriction, suggesting that stimulus-driven properties of the occluder, including luminance-related components (PLR), may have contributed to the effect. Although the present task was not designed as a standard pupillary light-reflex paradigm, the reduced constriction observed in patients is broadly consistent with previous reports of blunted pupil constriction during the pupillary light-reflex in schizophrenia (Rubin & Barry, 1976; Fattal et al., 2022). Differences between Acute and Chronic patients were small and inconsistent, suggesting that the pupil effect may reflect a broader alteration in task-evoked physiological responsivity rather than a clearly state-specific abnormality. This is also consistent with earlier pupillographic work showing no robust differentiation between acute and chronic schizophrenia groups across pupillary light-response measures (Hakerem et al., 1964).

Group comparison of late pupil constriction showed significantly reduced constriction in patients relative to controls. Reduced constriction was associated with greater negative symptom severity in the patient-by-session analysis (Figure 6d), whereas the patient-mean association was in the same direction but did not reach conventional significance. This finding is broadly consistent with previous reports linking blunted pupillary constriction to negative symptom severity in schizophrenia (Fattal et al., 2022) but should be interpreted cautiously given its analysis-dependent nature and the multiple physiological and medication-related factors that influence pupil size. In particular, medication effects on autonomic control of pupil size cannot be excluded. Thus, pupillary dynamics in the present task should be interpreted as a complementary physiological measure.

### Transdiagnostic oculomotor profiling

The present findings support a transdiagnostic view of oculomotor function. Rather than treating pursuit abnormalities as diagnosis-specific markers, this paradigm distinguishes components such as smooth tracking, gaze–target alignment, predictive tracking during occlusion, and pupil responsivity. Using the same paradigm in traumatic brain injury (TBI) (Shani et al., in press), we found broad impairments across pursuit initiation, saccadic pursuit, pupil responses, vergence, and occlusion-related tracking, which were associated with current functional status rather than initial injury severity. In contrast, psychosis showed a more selective profile: pursuit initiation and overall gain were relatively preserved, whereas tracking deviation and occlusion-related deviation were abnormal and associated with positive symptoms. Thus, the same task may identify distinct profiles across disorders—broad oculomotor disruption following diffuse neurological injury versus more selective impairment of gaze–target alignment and internally guided tracking in psychosis. This does not imply shared pathophysiology, but suggests that occlusion-related pursuit can serve as a dimensional transdiagnostic assay of sensorimotor prediction and control.

### Alternative explanations, medication effects, and limitations

Several alternative explanations should be considered. Reduced attention or arousal could contribute to smooth pursuit abnormalities in psychosis. However, the present pattern is unlikely to reflect generalized task disengagement alone. If global disengagement were the primary driver, broader impairments would be expected across basic oculomotor measures. Instead, full pursuit gain, initial catch-up saccade latency, and initial catch-up saccade amplitude were largely preserved, whereas abnormalities emerged in more specific measures of tracking deviation, gaze–target alignment, and occlusion-related deviation. Reduced pupil constriction also cannot by itself establish reduced attention, because pupil size is influenced by luminance-driven responses, autonomic regulation, fatigue, medication, and other physiological factors. Thus, generalized disengagement is unlikely to explain the full pattern, although more selective effects of attention or arousal on internally guided tracking cannot be excluded.

Medication is another important limitation. All patients were medicated as part of routine clinical care, and medication effects cannot be disentangled from illness-related mechanisms in the present study. Antipsychotic medication can influence oculomotor function, potentially through psychomotor slowing, sedation, or effects on pursuit gain (Sweeney et al., 1994; Hutton et al., 2001; Reilly et al., 2008). However, medication effects alone are unlikely to account for the full pattern of findings. Smooth pursuit deficits have been reported in unmedicated or neuroleptic-naïve patients and in unaffected first-degree relatives of individuals with schizophrenia, suggesting that such abnormalities cannot be attributed solely to pharmacological treatment (Holzman et al., 1973; Sweeney et al., 1994; Thaker et al., 1999; Calkins et al., 2004; Calkins et al., 2008). Moreover, several effects were stronger in Acute patients despite medication in both patient groups, suggesting a contribution of current clinical state. Nevertheless, medication type, dose, duration, and polypharmacy remain important potential confounds. Future studies including unmedicated patients, medication-stratified samples, or quantitative antipsychotic-dose analyses will be needed to clarify these effects.

Additional limitations should also be noted. The sample size was modest, particularly for comparisons between Acute and Chronic patient groups, and replication in larger samples will be important. Patients were tested across one to four sessions, whereas healthy controls were tested once; although this design allowed examination of clinical fluctuation within the patient group, it did not fully match groups for repeated testing. Healthy controls were recruited primarily from hospital staff and affiliated academic trainees, which may limit generalizability. Finally, the use of a remote 90-Hz Tobii eye tracker under head-free bedside conditions improved clinical feasibility but limits fine temporal interpretation, particularly for single-trial saccade timing. The main findings were based primarily on spatial and trial-averaged measures, but future studies using higher-resolution eye tracking and more controlled head stabilization would help validate and refine these findings.

## Conclusions

The present findings suggest that smooth pursuit abnormalities in psychosis are expressed differently across pursuit measures and clinical states. During continuous tracking, basic pursuit-related measures were largely preserved, whereas smooth tracking gain was reduced primarily in Acute patients and tracking deviation was altered in both patient groups; tracking deviation and occlusion-related deviation were also associated with positive symptom severity. Structured occlusion extended the conventional pursuit paradigm by increasing the need to anticipate and maintain the expected target trajectory, revealing differences between patient groups that were less apparent during continuous pursuit.

## Data Availability

The data are not publicly available at the preprint stage. De-identified data supporting the findings of this study will be made available upon publication in a peer-reviewed journal, subject to ethical and institutional approvals.

## Notes

### Competing Interest Statement

The authors have declared no competing interest.

### Author Declarations

The study was approved by the Kaplan Medical Center Helsinki Committee.

